# Predicted loss of function alleles in Bassoon (BSN) are associated with obesity

**DOI:** 10.1101/2023.02.19.23285978

**Authors:** Na Zhu, Charles A. LeDuc, Ilene Fennoy, Blandine Laferrère, Claudia A. Doege, Yufeng Shen, Wendy K. Chung, Rudolph L. Leibel

**Author notes:** Co-senior/corresponding authors. Co-first authors (Na Zhu, Charles A. LeDuc).

## Abstract

Bassoon (*BSN*) is a component of a hetero-dimeric presynaptic cytomatrix protein that orchestrates neurotransmitter release with Piccolo (*PCLO*) from glutamatergic neurons throughout the brain. Heterozygous missense variants in *BSN* have previously been associated with neurodegenerative disorders in humans. We performed an exome-wide association analysis of ultra-rare variants in about 140,000 unrelated individuals from the UK Biobank to search for new genes associated with obesity. We found that rare heterozygous predicted loss of function (pLoF) variants in *BSN* are associated with higher BMI with log10-p value of 11.78 in the UK biobank cohort. The association was replicated in the All of Us whole genome sequencing data. Additionally, we have identified two individuals (one of whom has a *de novo* variant) with a heterozygous pLoF variant in a cohort of early onset or extreme obesity at Columbia University. Like the individuals identified in the UKBB and All of us Cohorts, these individuals have no history of neurobehavioral or cognitive disability. Heterozygosity for pLoF *BSN* variants constitutes a new etiology for obesity.

## Introduction

By 2030 it is estimated that roughly 50% of adults in the United States will have obesity, with 25% having severe obesity (1). The prevalence of obesity in U.S. adults has increased from 30.5 to 41.9% from 1999-2000; the prevalence of severe obesity has increased from 4.7 to 9.2%. Approximately 18% of U.S. children currently have obesity (2). Variously estimated, the risk of obesity is 30-50% heritable (3-6). Changes in the underlying genetics cannot be responsible for such changes in the prevalence of obesity over such a short period of time; however, the propensity to gain weight in an environment with ready access to food is largely genetic (7). Genome wide association studies have identified many common variants associated with body weight regulation (8-10). More recently, polygenic risk scores aggregating large numbers genetic variants, each with small contributions to energy homeostasis can be used to predict obesity deciles in some genetic ancestries (11). However, the genetic attributable risk for obesity remains modest at ∼3% (12, 13). Exome sequencing of large numbers of individuals has accelerated the discovery of rare genetic contributors to quantitative phenotypes such as height (14, 15), celiac disease (16), and dyslipidemia (17, 18). In many instances the precise mechanistically functional relevance of these associated genetic variants remains unknown.

Recent advances in the treatment of obesity (19) and hyperlipidemia (20) have used human genetics to identify genes contributing to extreme phenotypes to understand biology and molecular mechanisms and develop novel interventions. The advent of large-scale exome/genome sequencing in the United Kingdom Biobank (UKBB) and All of US has extended the ability to assess rare variants at large scale in addition to prior methods of assessing common variants in GWAS. In the current study we combine the power of exome sequence-based analysis of an extreme obesity cohort recruited at Columbia University with data from the UKBB and All of Us. We report the association of predicted loss of function (pLoF) alleles in the gene *BSN* with body mass index (BMI).

## Methods and Materials

### Methods

#### Columbia Cohort

The Columbia University Extreme Obesity cohort was collected using protocols approved by the Institutional Review Boards at Columbia University Irving Medical Center (New York, NY) and The Rockefeller University (New York, NY). The cohort consists of 1598 individuals from 903 families. Obesity was defined as described below. Of the 903 families, 122 constitute affected child/parent trios. The remaining 781 families have 1372 affected (890 females and 482 males) and 226 unaffected family members. Cohort details have been described previously (21, 22).

Approximately half of the probands were pediatric (either recruitment or obesity onset age younger than 19 years old with 674 participants having a BMI Z score >=2; average age at enrollment 6.6 +/- 3.6 years) and half adults (obesity onset or recruitment age at least 19 years old with 698 adults with BMI >=30 kg/m^2^; average age 51.5 +/- 12.0 years) (Table 1). Samples were exome sequenced using xGen and SeqCap VCRome Capture. Greater than 99% of samples had depth of coverage > 10 in 80% of target regions.

**Table 1.**
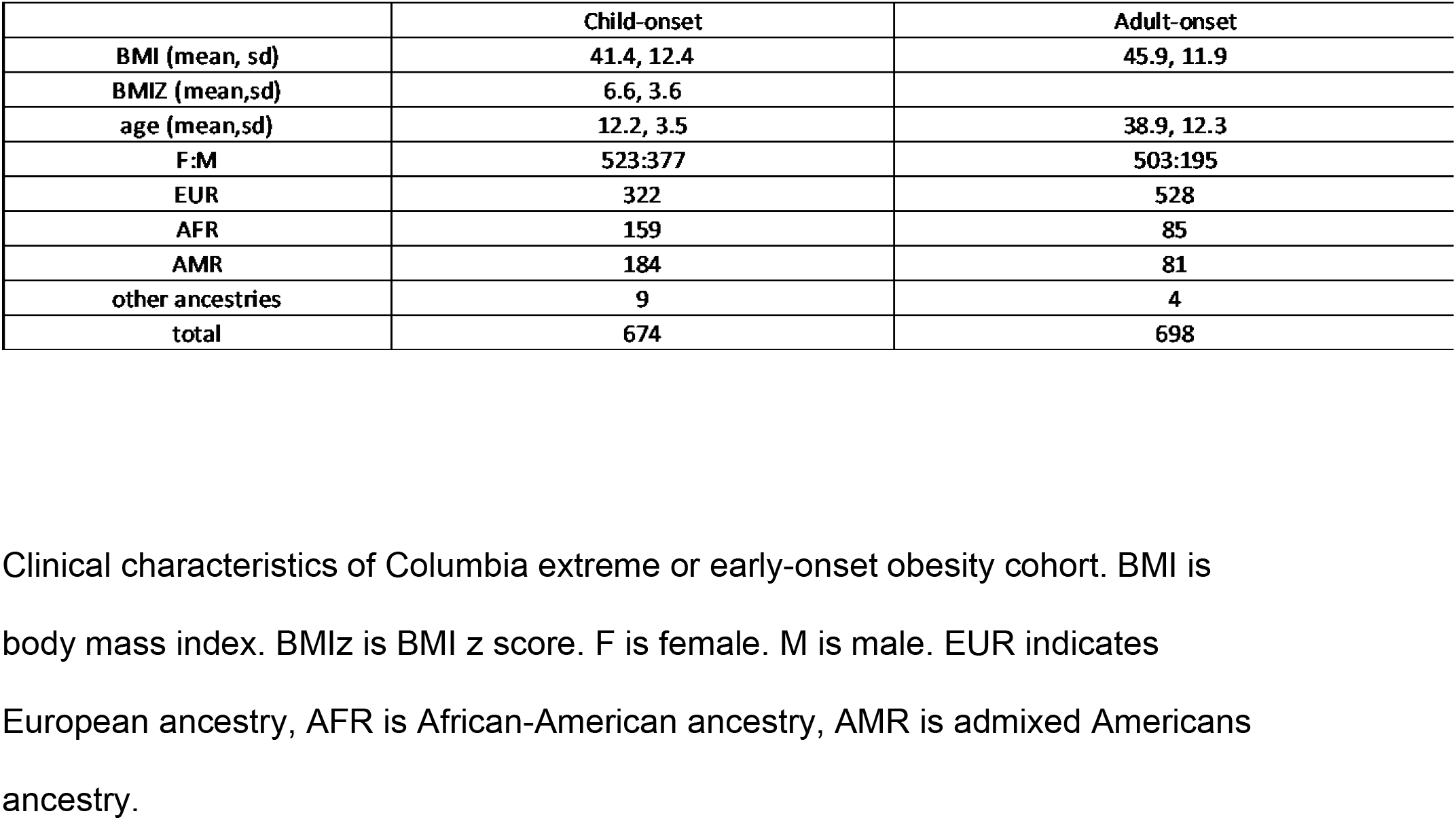

Controls were the unrelated parents (without autism) from the Simons Powering Autism Research for Knowledge (SPARK) study and were exome sequenced using the XGEN-Capture (23).

#### UKBB Cohort

For this analysis, we included 200,643 UK biobank cases (24). The average age of this cohort is 56.4 +/- 8.1 years; mean BMI of 27.3 +/- 4.8 kg/m^2^; 55.1% female (Table 2).

**Table 2.** Summary of United Kingdom Biobank subjects.

| Table 2 UKBB cohort summary |  |
| --- | --- |
| Overall Cohort |  |
| BMI (mean, sd) | 27.3, 4.8 |
| age (mean, sd) | 56.5, 8.1 |
| F:M | 110476:90153 |
| EUR ancestry | 167,246 |
| ---removed for relatedness | 4,878 |
| ---had cancer | 16,711 |
| ---had eating disorder | 112 |
| ---cancer and eating disorder | 18 |
| Cohort Included in study |  |
| EUR no cancer, no eating disorder | 145,103 |
| F:M | 78103:67000 |
| BMI (mean, sd) | 27.5, 4.7 |
| age (mean, sd) | 56.6 |
| Correlation between age and BMI =0.048, significant |  |
| correlation between age and Sex =0.082, significant |  |

The UKBB cohort use in the analysis. Samples that were coded with, cancer, eating disorders, or both were removed from the cohort prior to analysis.
Relatedness was estimated using plink King, when sample pairs had a relatedness greater than 0.12 (second degree relative or closer) the sample that had more relatedness to the cohort was excluded.

#### All of Us data

The current release (June 2022) of the All of Us data includes whole genome sequencing for 98,622 individuals (58,190 females and 38,290 males). The average age of this cohort is 52.6 +/- 16.9 years; mean BMI is 30.9 +/- 9.0 kg/m^2^.

### Bioinformatic analysis of exome or genome sequencing data

#### Columbia cohort

Paired-end reads were mapped and aligned to the human reference genome (version GRCh38/hg38, accession GCA 000001405.15) using BWA-MEM (25). Picard v1.93 MarkDuplicates (http://broadinstitute.github.io/picard/) was used to identify and flag PCR duplicates and GATK v4.1 HaplotypeCaller (26) in Reference Confidence Model mode to generate individual-level gVCF files from the aligned sequence data. We performed joint calling of variants from the obesity cohorts using GATK variant caller.

#### Ancestry prediction and relatedness check

We predicted the ancestry and relatedness in the Columbia cohort using Peddy (27). Relatedness prediction in the UKBB samples, due to the large sample size, was completed with plink King (28). When pairs of samples shared second degree relationship or closer (a kinship coefficient greater than 0.12 in King or 0.25 in Peddy), the sample with greater relatedness to the cohort was excluded.

#### Variant annotation

We used the Ensembl Variant Effect Predictor (VEP, Ensemble 93) (29) to annotate variant function and ANNOVAR (30) to aggregate variant population frequencies and for *in silico* predictions of deleteriousness. Rare variants were defined by a population frequency < 10^−4^ in gnomAD WES and WGS (31). Deleterious variants were defined as <u>predicted loss of function</u> (pLoF: including premature stop-gain, frameshift indels, canonical splicing variants and multi-exon deletions) or <u>predicted damaging</u> missense (Dmis) based on REVEL (32) score thresholds. The same pipeline was used for Columbia, UKBB, and All of US variant annotation.

### Statistical analysis

#### Columbia cohort

We tested the single variant association with obesity using the exact binomial test in the unrelated European participants. To identify novel risk genes for obesity in the Columbia cohort, we performed a rare variant gene burden test using the binomial test in unrelated European participants. When there were multiple individuals with obesity in a family, we defined the most severely affected as the proband (defined as the child with the highest Z-score or the adult with the highest BMI if there were only adults in the family).

A gene-based case-control association test was performed on 483 unrelated cases and 11,101 unrelated SPARK non-autism parents as population controls by comparing the frequency of rare deleterious variants in obese cases with SPARK controls. To minimize false positive variant calls and reduce batch effect, we applied additional heuristic filters in cases and controls by the following exclusion criteria (a variant was excluded if any one condition was met):

Variants were filtered out if any of these exclusion criteria were met: (a) cohort allele frequency was > 0.01; (b) the variant was not uniquely mappable; (c) genotype allelic fraction was < 0.2; (d) the variant was shared in multiple cases (alternate allele count > 4) with at least half of the cases with low quality calls (allelic fraction < 0.35); (e) less than 90% of individuals (cases and controls) have ≥10x depth of average of the variant site. (f) All variants in SPARK parents were required to pass the deep variant test (33, 34). (g) All single nucleotide variants (SNVs) were high quality calls defined by GATK VQSLOD > -3 in the case cohort.

To assess the overall degree of batch effects, we compared the rare synonymous variant frequencies between cases and controls, testing the assumption that most rare synonymous variants do not have effects on obesity. A gene level burden test QQ plot for synonymous variants shows deflation with lambda=0.75 due to the limited case sample size resulting in genes that had no variants in the cases. Nevertheless, observed p-values were consistent with the expected p-value in the testable genes (Sup Figure 1).

To identify obesity-risk genes, we tested the deleterious variant burden (pLoF or Dmis) in each protein-coding gene in cases compared to controls using an exact binomial test. REVEL scores were used to predict the deleteriousness of missense variants. We performed 20 association tests for each gene, including pLoF only, Dmis only and Dmis + pLoF where Dmis was defined using 5 different REVEL score cutoffs (0.15 to 0.95 by 0.2).

#### UKBB cohort

After excluding related individuals and individuals with a history of cancer or eating disorder, 144,496 unrelated European individuals were selected for quantitative trait (BMI) association analysis (31, 35). We collapsed rare variants based on allele frequency and predicted variant deleteriousness. The variants were partitioned into cohort frequency <10^−4^ and singleton population allele frequency groups as well as 10 variant functional groups. The variant functional groups were missense variants with REVEL >=x, with x ranging from 0.15 to 0.95 in 0.2 increments with or without pLoF variants (10 groups). Genes with less than 15 heterozygotes in a test group were removed. The significance threshold was set at (0.05/ (20*20,000)). We then tested the quantitative BMI for the 144K UKBB individuals using REGENIE (36), which accounts for relatedness, population structure and polygenicity. We included age, Townsend deprivation index at recruitment, smoking /alcohol status, sex, the first 8 principal components, and genetic heterozygosity as covariates. REGENIE resolved the gene-based association tests in the large UKBB dataset with no inflation or deflation in the synonymous variants with the gene-based tests (Sup Figure 2a). The type I error rate was well controlled for pLoF and Dmis variants in gene-based tests, showing minor inflation in the QQ plot (Sup Figure 2b).

Finally, we ran a meta-analysis using Fisher ‘s method (https://cran.r-project.org/web/packages/metap/index.html) for UKBB and Columbia samples with the same defined variant groups. We defined the threshold for genome-wide significance by Bonferroni correction for multiple testing (n=20,000*20, threshold p-value=1.3e-7) (workflow shown in Figure 1).

**Figure 1.**
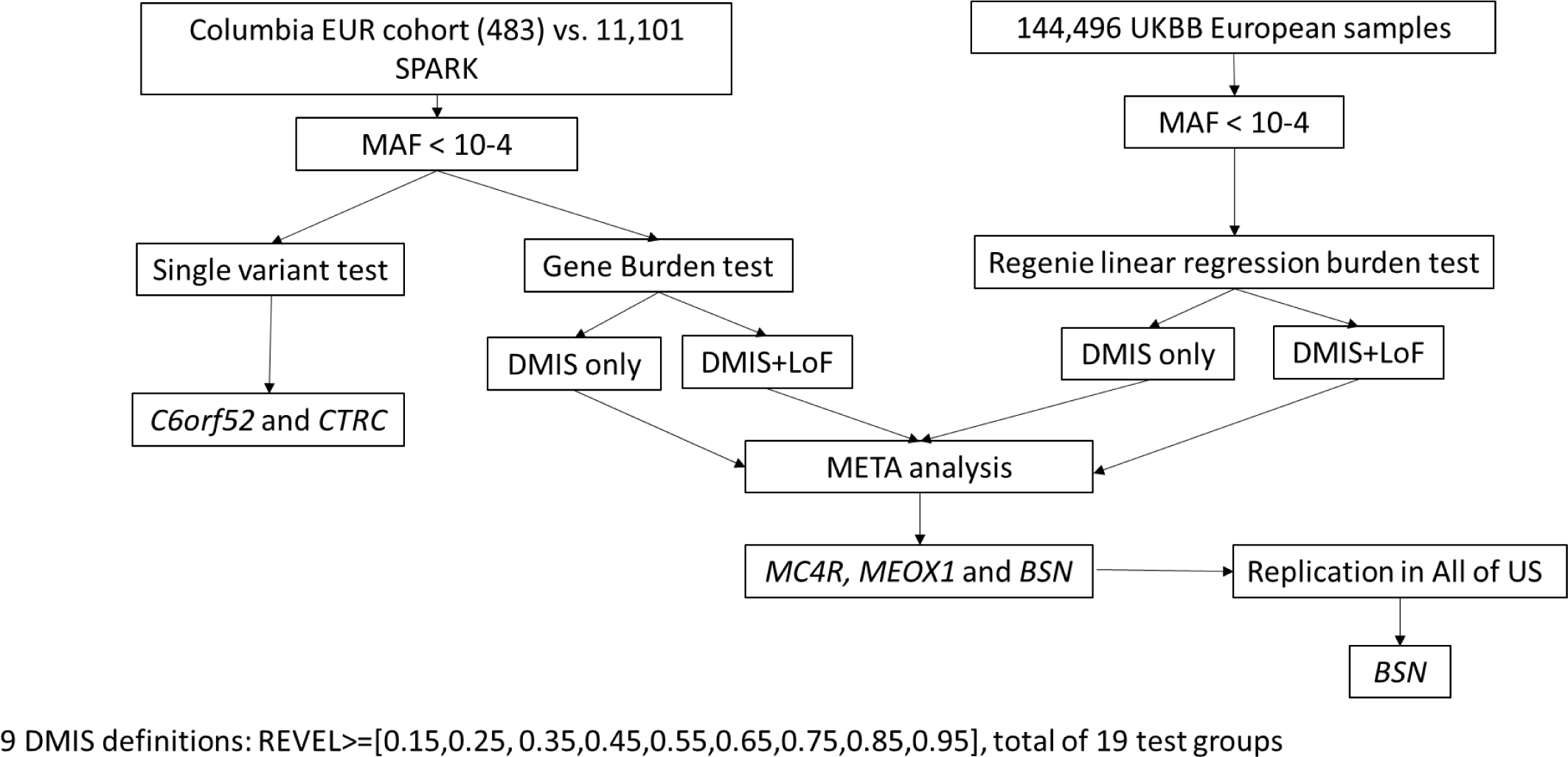
Summary of workflow Workflow summary. Only rare (MAF <10^−4^) variants are filtered. For the Columbia European cohort, all rare variants are put through both a single variant test and a gene burden test. For the UKBB variants, all rare variants are tested using the regenie linear regression burden test. Genes with 19 different Dmis definitions are compared in a Meta analysis that integrates both datasets. Replication in the three genes that reached significance were attempted in the All of Us dataset, with *BSN* replicating.

#### All of Us

To attempt to replicate findings from the UKBB analysis, we ran a linear regression on the 48,722 European ancestry individuals from the All of Us dataset using their provided cloud-based research platform to test the association between BMI and *BSN* and *MEOX1* deleterious variants using age, sex, deprivation index and median income as covariates.

## Results

In the single variant association tests, we identified two exome-wide significant single nucleotide variants (SNVs) in the Columbia cohort. rs887287256 is a c.C477A: p. Asp159Glu variant in *C6ORF52* (NM_001388310.1). The Columbia cohort had six unrelated European individuals with obesity who were heterozygous, and no heterozygotes or homozygotes in 11,101 SPARK controls (-log10p, 8.27, RR=276). rs202058123, a c.G649A: p.Gly217Ser SNV in *CTRC* was present in five heterozygotes with obesity in the Columbia cohort and one of the SPARK controls (-log10p 6.1, RR =114) (Supplementary Table 1-3). We performed segregation analysis for those Columbia families with available family members (Supplementary Figure 3 and 4). All heterozygotes had obesity, but not all individuals with obesity in the family had the relevant variant. However, neither variant association was replicated when tested using the UKBB data.

We performed gene-based burden tests with 20 groups tested for each gene. Twelve tests in three unique genes (*MC4R, BSN*, and *MEOX1*) passed Bonferroni corrected significance (-log10 p-value >=6.9) in the combined association tests. Using a false discovery rate < 0.1, the most significant gene-variant sets are listed in Table 3, Supplemental Table 4 and 5.

**Table 3.**
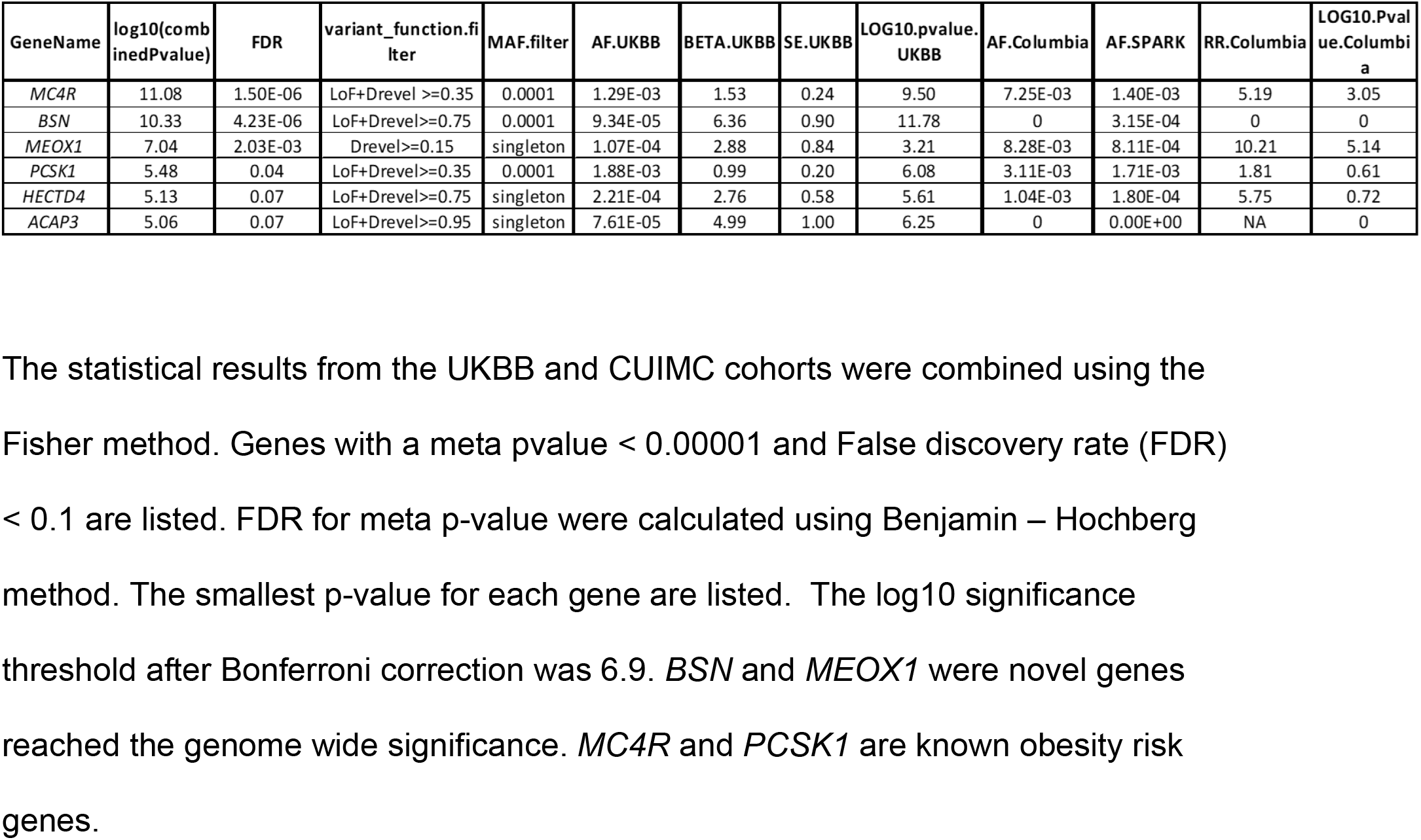
Meta analysis for UKBB REGENIE linear regression and Columbia binary burden test

| GeneName | log10(combinedPvalue) | FDR | variant_function.filter | MAF.filter | AF.UKBB | BETA.UKBB | SE.UKBB | LOG10.pvalue.UKBB | AF.Columbia | AF.SPARK | RR.Columbia | LOG10.Pvalue.Columbia |
| --- | --- | --- | --- | --- | --- | --- | --- | --- | --- | --- | --- | --- |
| *MC4R* | 11.08 | 1.50E-06 | LoF+Drevel >=0.35 | 0.0001 | 1.29E-03 | 1.53 | 0.24 | 9.50 | 7.25E-03 | 1.40E-03 | 5.19 | 3.05 |
| *BSN* | 10.33 | 4.23E-06 | LoF+Drevel>=0.75 | 0.0001 | 9.34E-05 | 6.36 | 0.90 | 11.78 | 0 | 3.15E-04 | 0 | 0 |
| *MEOX1* | 7.04 | 2.03E-03 | Drevel>=0.15 | singleton | 1.07E-04 | 2.88 | 0.84 | 3.21 | 8.28E-03 | 8.11E-04 | 10.21 | 5.14 |
| *PCSK1* | 5.48 | 0.04 | LoF+Drevel>=0.35 | 0.0001 | 1.88E-03 | 0.99 | 0.20 | 6.08 | 3.11E-03 | 1.71E-03 | 1.81 | 0.61 |
| *HECTD4* | 5.13 | 0.07 | LoF+Drevel>=0.75 | singleton | 2.21E-04 | 2.76 | 0.58 | 5.61 | 1.04E-03 | 1.80E-04 | 5.75 | 0.72 |
| *ACAP3* | 5.06 | 0.07 | LoF+Drevel>=0.95 | singleton | 7.61E-05 | 4.99 | 1.00 | 6.25 | 0 | 0.00E+00 | NA | 0 |

The statistical results from the UKBB and CUIMC cohorts were combined using the Fisher method. Genes with a meta pvalue < 0.00001 and False discovery rate (FDR) < 0.1 are listed. FDR for meta p-value were calculated using Benjamin – Hochberg method. The smallest p-value for each gene are listed. The log10 significance threshold after Bonferroni correction was 6.9. *BSN* and *MEOX1* were novel genes reached the genome wide significance. *MC4R* and *PCSK1* are known obesity risk genes.

Limiting the analysis to pLoF and Dmis variants with REVEL score >=0.25, the association test was genome-wide significant for MC4R, with a BMI effect size beta in UKBB of 1.4 kg/m^2^ and relative risk for obesity of 5.03 in the Columbia cohort. The UKBB and Columbia heterozygotes are listed in Supplemental Table 6 and 7 and Supplemental Figure 5. Effect size was estimated with a linear regression test run on individual variants.

The combined (Columbia and UKBB) p-value (-log10P:10.33) for BSN reached genome-wide significance. This signal is primarily driven by the UKBB data since pLoF and Dmis variants with REVEL score >=0.75 are extremely rare (AF in UKBB was 9.3e-05) and few in number in the smaller Columbia cohort. The UKBB data alone have a strong signal with a BMI effect size beta of 6.21 and -log10p of 11.78. No positive effect size is observed in other missense groups. All heterozygous predicted deleterious variants in UKBB are listed in Supplemental Table 8. Figure 2a shows the BMI distribution of BSN predicted deleterious heterozygotes compared to the overall UKBB population (Kolmogorov-Smirnov pvalue 1.4e-05).

**Figure 2.**
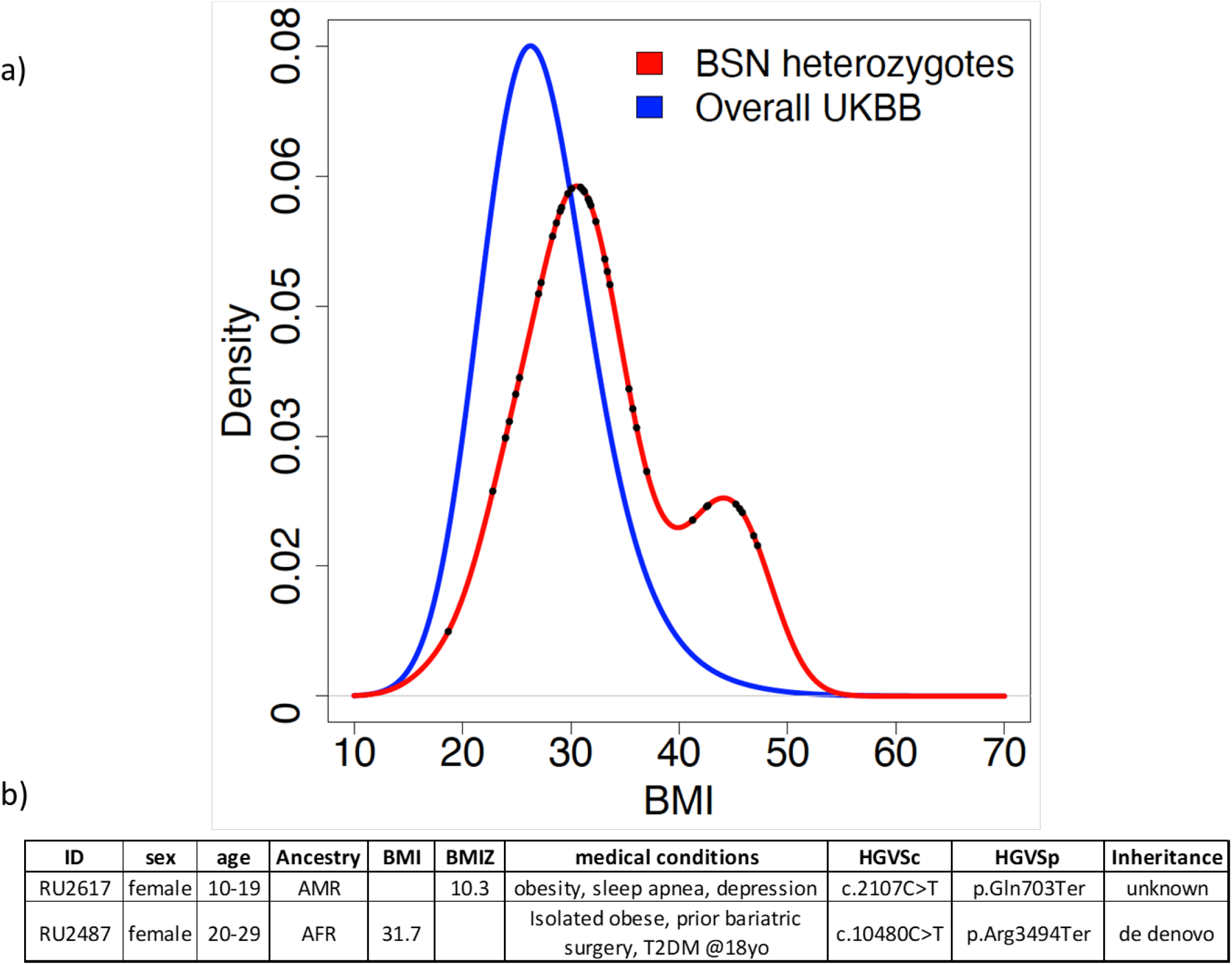
BMI density distribution. a) The BMI density distribution for pLoF *BSN* heterozygotes is shifted to a higher BMI than the overall UKBB cohort. A.) There is a bi-modal distribution for the *BSN* pLoF heterozygotes. The distribution difference between overall cohort and UKBB heterozygotes was tested using the Kolmogorow-Smirnov method (p=1.4e-05). The dots in the blue curve represent the *BSN* predicted deleterious variants samples ‘ BMI. b) Phenotype of *BSN* pLoF heterozygotes in the Columbia cohort.

Two heterozygous pLoF *BSN* alleles were identified in the Columbia cohort (Figure 2b). Study IDs are known only to the study staff. RU2487 is heterozygous for a *de novo* p.Gln703X allele in *BSN*. At the time of the last assessment, she was a Latina woman in her 20 ‘s with a history of severe obesity and type 2 diabetes mellitus diagnosed as a teen at which time her HbA1c was 7.4%. She was amenorrheic and had extensive acanthosis nigricans, dyslipidemia, hypothyroidism, and hyperandrogenism. Her maximal weight was 113 kg. She had gastric bypass surgery for weight loss in her 20 ‘s. Immediately prior to bariatric surgery, her BMI was 39.7 kg/m^2^. Her oral glucose tolerance test prior to bariatric surgery showed euglycemic hyperinsulinemia. Her nadir body weight after surgery was 77 kg; 2 years post-operatively she weighed 101 kg. She reports frequently feeling very hungry. She is a college graduate with no academic or cognitive difficulties nor history of psychiatric diagnoses. She has no family history of obesity or type 2 diabetes.

RU2617 is an African American female heterozygous for a p.R3494X variant in *BSN*; the allele was not inherited from the only parent available for genetic analysis. At the time of her initial evaluation, the patient was a teen with body weight of 162 kg and height of 160.9 cm (BMI=62.6 kg/m^2^). Her waist circumference was 158 cm. She had no history of irregular periods. She had obstructive sleep apnea requiring continuous positive airway pressure. She initially had a normal glucose tolerance test with normal fasting glucose and HbA1c = 6.3%; however, she subsequently developed impaired fasting blood glucose of 105 mg/dl with persistently elevated HbA1c. She had laparoscopic adjustable gastric banding as a teen. At 3 years post operatively, her weight had declined to 134.2 kg and her height had increased to 163 cm (BMI of 50.5 kg/m^2^). HbA1c normalized to 5.2%.

The association of *MEOX1* with BMI was genome-wide significant (-log10P: 7.04) in the combined analysis of the UKBB and Columbia cohorts. In the Columbia cohort, deleterious missense variants (REVEL >=0.15) were 10.2 times more frequent than in the SPARK participants. The singleton deleterious variants in *MEOX1* were marginally significantly associated with BMI in UKBB (-log10P 3.21, beta 0.84). The majority of singleton predicted deleterious *MEOX1* variants in the UKBB were associated with a higher BMI. The BMI in individuals with heterozygous *MEOX1* deleterious missense variants was significantly higher than the overall UKBB (p value 0.03 using the Kolmogorov-Smirnov test). (Figure 3a and Supplemental Table 9). In the Columbia cohort, *MEOX1* predicted Dmis variants (Table 4) were enriched in the pediatric-onset compared with the adult-onset obesity cases. There were 7 heterozygotes out of 262 unrelated European ancestry obese children and 2 heterozygotes out of 362 unrelated European ancestry obese adults (p=1.2e-6 with a relative risk of 16.5 for the pediatric-onset group and p= 0.13 with a relative risk of 3.4 in adult-onset group). For the *MEOX1* individual variants (Table 4 and Figure 3b), missense variant p.R213H (CADD score 27.5 and REVEL 0.926, indicating likely deleterious) was observed in 3 pediatric-onset and 1 severe adult-onset individuals in the Columbia cohort; there were none in SPARK participants. In the UKBB there were two heterozygote participants with p.R213H variants with BMI 26.4 and 29.9 kg/m2. Across the combined TOPMED and gnomAD databases p.R213H was observed only once. The missense variant p.R184Q (CADD score 28.1, REVEL score 0.662) was observed in three pediatric-onset individuals in the Columbia cohort and twice in the SPARK participants. In the UKBB, there were 10 heterozygotes: one had obesity, seven had overweight and two had normal BMI. The population frequency of the p.R184Q variant is 8.5e-05 in gnomAD and 8e-05 in TOPMED. Segregation analysis for the Columbia *MEOX1* heterozygotes showed that all the heterozygotes in those families had obesity (Figure 4).

**Figure 3.**
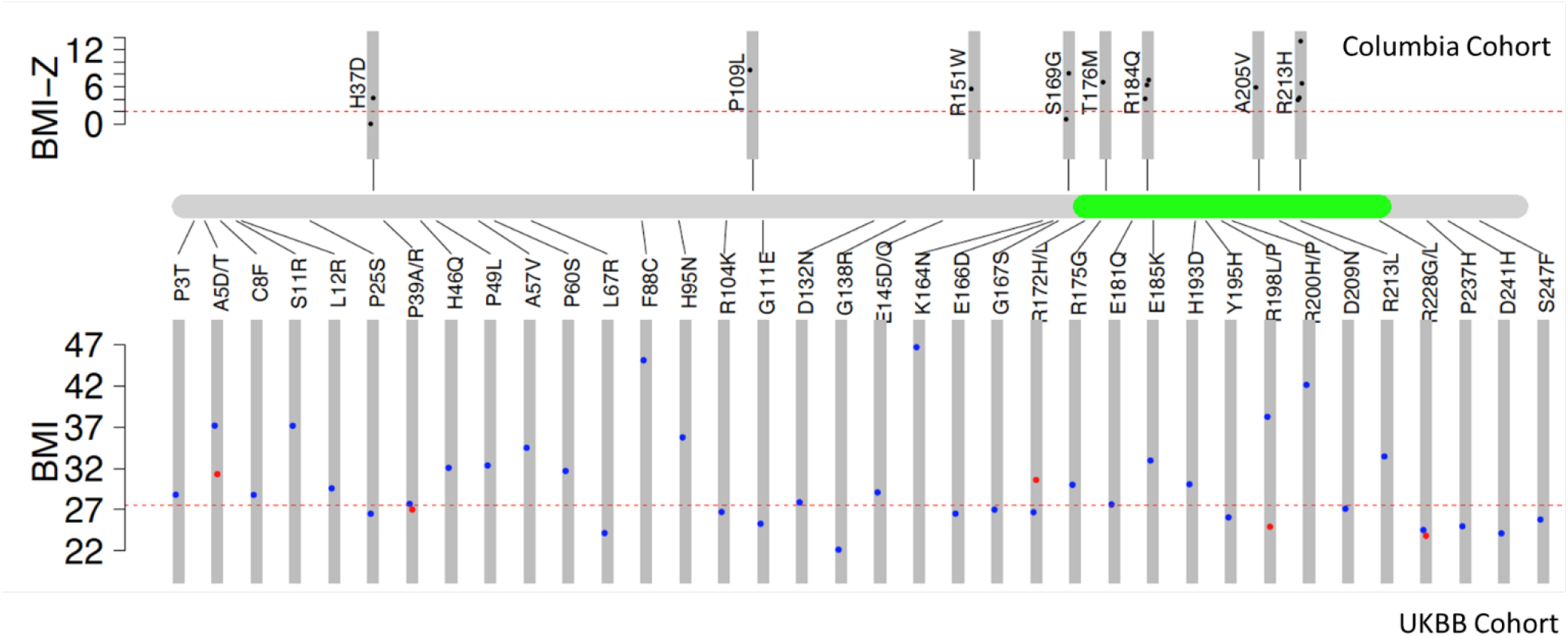
Lollipop plot for *MEOX1*. The upper panel shows BMI-Z distribution for rare deleterious variants (population frequency < 10-4 and revel score>=0.15) in the Columbia cohort. BMI-Z for adult samples was the normalized BMI score using UKBB mean and standard deviation and BMI-Z for the child was the raw BMI-Z score. The lower panel shows the singleton deleterious variants in the UKBB. The BMI distribution difference between *MEOX1* singleton deleterious variants carriers with the overall UKBB cohort is 0.03.

**Figure 4.**
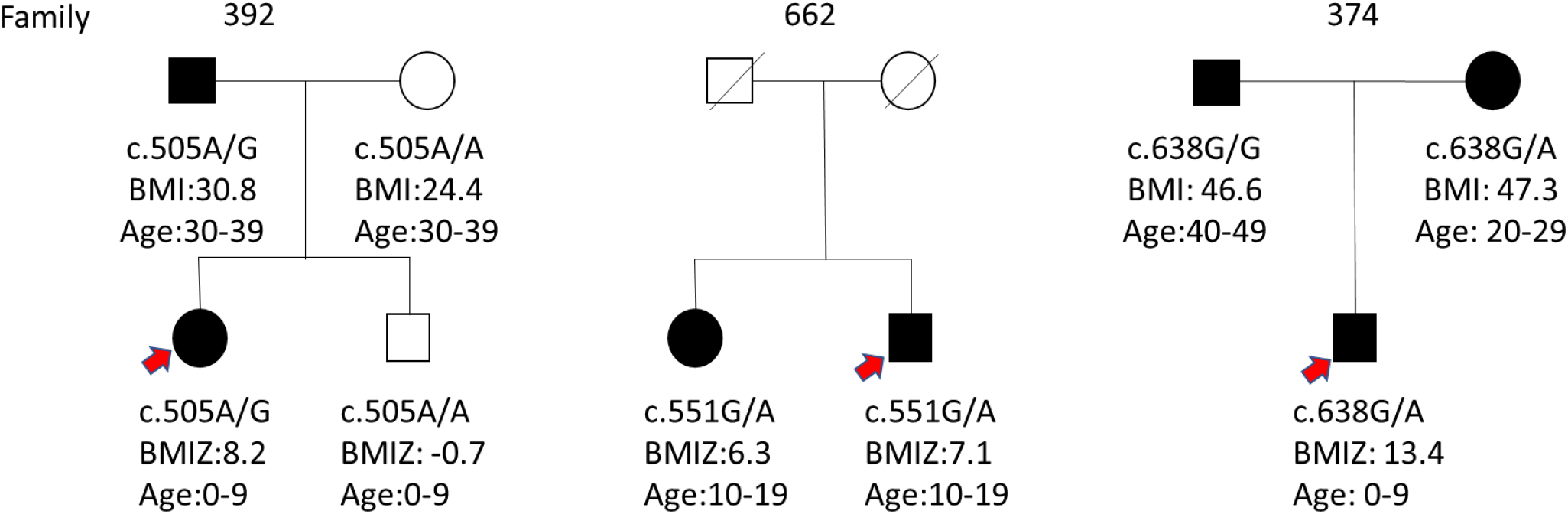
*MEOX1* segregation Columbia *MEOX1* heterozygous pedigrees. Shaded symbol represents obese person, red arrow indicates the proband in the family. BMI or BMIZ are indicated under the symbols.

**Table 4.**
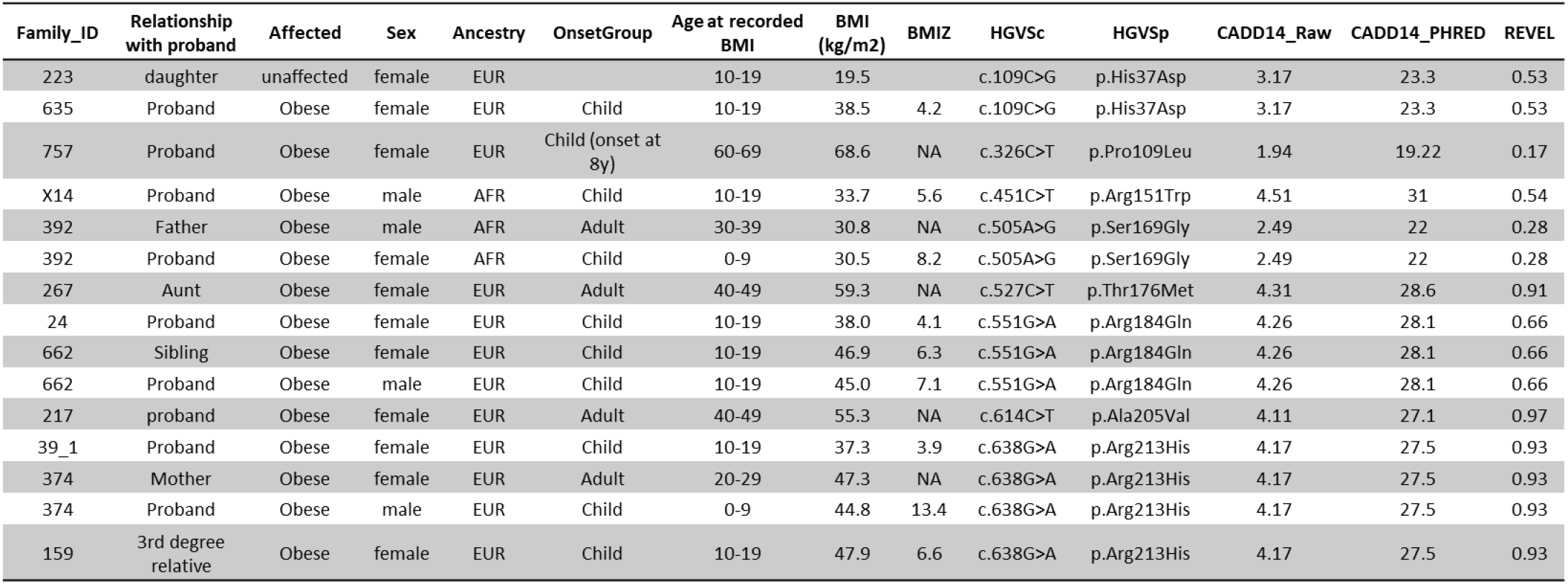
Predicted deleterious variants in *MEOX1* in the Columbia cohort. Phenotype, variant, predicted variant severity, and ancestry of the *MEOX1* heterozygotes in Columbia cohort.

### Association of BMI-correlated traits in *BSN*

The association between BSN and the traits correlated with BMI tested using REGENIE (Table 5) showed arm, leg and trunk fat mass and leg fat-free mass and leg predicted mass reached genome-wide significance. We also tested the association between *BSN* and ICD10 diagnoses (Supplemental Table 10) using the binomial test. No diagnosis was significantly associated with *BSN* after correction for multiple testing.

### Replication analysis using All of Us data

We identified *BSN* and *MEOX1* heterozygotes in the All of Us cohort. To date, there are 98,622 subjects for whom both whole genome sequencing and clinical data are available. Half of the participants (47,897) are unrelated and of European ancestry. For each participant, we used the highest recorded BMI, giving a cohort average BMI of 30.1 +/- 7.8 kg/m^2^. In the cohort, 12 European individuals were heterozygous for *BSN* pLoF variants, with an average BMI of 37.0 +/- 5.7 kg/m^2^. Using sex, age, income, and deprivation index as covariates, we tested the association between BMI and *BSN* genotype using linear regression and found a significant association (p-value=0.0075, beta=6.27). Additionally, we identified an additional six *BSN* pLoF heterozygotes among the non-European participants (mean BMI 31.5 (SD = 8.5 kg/m^2^); BMI range = 22-45; 3/6 with BMI >30.0; Supplemental Table 11). Thus, the *BSN* obesity association observed in the UKBB and Columbia cohorts was replicated in the All of Us cohort. *MEOX1* predicted deleterious variants were not associated with higher BMI in All of Us (pvalue=0.47, beta=0.57).

## Discussion

We have identified a gene, *BSN*, for which we have demonstrated an association of rare pLoF variants with obesity in two independent large cohorts: the UKBB and All of Us. Additionally, we identified extremely obese individuals in the Columbia cohort of extreme obesity, including an individual with extreme, early onset obesity associated with a *de novo* pLoF allele. There is no evidence that these variants are associated with intellectual disability or cognitive impairment, including direct assessment of two individuals in the Columbia cohort. A second gene, *MEOX*, was identified with predicted Dmis variants associated with obesity in the UKBB and Columbia cohorts, but this finding was not replicated in the All of Us cohort.

*BSN* (bassoon) is expressed primarily in the brain (including embryonic and adult brain regions that impact feeding behavior (37)), inner hair cell ribbons, and the retina of mammals. Bassoon is a presynaptic scaffold protein localized in the cytomatrix at the active zone (CAZ) where it functions to orchestrate neurotransmitter release. Bassoon participates in the formation of Golgi-derived Piccolo-Bassoon transport vesicles that are axonally transported to newly formed synaptic contacts. Bassoon is associated with activity-dependent short- and long-term neuronal plasticity (38).

Bassoon is expressed during early neuronal differentiation, is selectively sorted into axons and is among the first proteins to arrive at nascent synapses (38). The release of neurotransmitters from the presynaptic terminal involves the active zone (AZ). The AZ includes an electron-dense protein meshwork, the presynaptic cytomatrix.

Bassoon is one of several scaffolding proteins (along with Piccolo (*PCLO*), *RIM, MUNC13*, and *ELKS*) within the presynaptic cytomatrix. *BSN* and *PCLO* are structurally related, interact, and are the largest active-zone-specific proteins. Unlike other the proteins in the AZ that are evolutionally conserved down to C. elegans, Piccolo and Bassoon are only found in vertebrates (39).

Mice homozygous for LoF *Bsn* alleles have reduced synaptic transmission that is primarily caused by the inactivation of a significant fraction of glutamatergic synapses. These mice have spontaneous epileptic seizures. Bassoon is not essential for synapse formation but is essential for regulated neurotransmitter release from a subset of glutamatergic synapses. (40). At the ultrastructural level, these inactive synapses cannot be distinguished from functional synapses. These homozygous Bassoon mutant mice have seizures with structural brain alterations including enlarged hippocampi and cerebral cortices (41). These animals are not obese.

Bassoon is involved in the maintenance of the integrity of AZ (42). Glutamatergic synapses from *Bsn* knockout mice exhibit enhanced short-term synaptic depression with a high percentage of silent synapses but have no gross structural defects (43), presumably due to the significant functional redundancy with Picolo. When both proteins are absent from glutamatergic synapses, the cells undergo synapse degeneration (44).

*BSN* was originally identified while attempting to identify expressed cerebellar transcripts in patients with multiple system atrophy, a rare progressive neurodegenerative disease characterized by cerebellar symptoms, parkinsonism, and autonomic dysfunction (45). This study did not find coding mutations in *BSN* but first identified *BSN* as a new transcript that they could clone from the cerebellum of these patients. *BSN* acts in concert with Parkin RBR E3 Ubiquitin Protein Ligase (PRKN) to control presynaptic autophagy and maintain homeostatic presynaptic proteostasis and synaptic vesicle turnover (46). Human heterozygous missense variants in *BSN* have been implicated in neurodevelopmental and neurodegenerative disorders including progressive supranuclear palsy-like syndrome, a rare neurodegenerative tauopathy (47).

We have implicated heterozygous pLoF variants in *BSN* as a new genetic etiology for human obesity that is not associated with adverse impact on cognition or other neurobehavioral phenotypes. The expression of *BSN* throughout the brain suggests that gene dosage could contribute to hyperphagia through both homeostatic and hedonic circuits (48). Additional detailed phenotypic assessment – ideally of individuals prior to the onset of obesity - will be required to assess this point. *BSN* is expressed in the synapses of glutamatergic neurons and hypothalamic neurons mechanistically tied to ingestive behaviors (43, 49-51). The valence of these effects is consistent with hyperphagic obesity conveyed by hypomorphic alleles.

## Data Availability

All data produced in the present study are available upon reasonable request to the authors

## Declarations

Competing interests: No author has any conflicts or competing interests rated to the manuscript. IRB: All studies were under the auspices of the Columbia University IRB “Molecular Genetic Analysis of Obesity and Non-Insulin Dependent Diabetes Mellitus” IRB #: AAAA4485 which expires on 5/1/23.

## Acknowledgements

We thank the participants who generously contributed to this work and their clinicians who referred them.

This work was supported by NIH grant NIDDK 52431 and the NY Nutrition and Obesity Research Center: P30DK26685.

## Notes

### Competing Interest Statement

The authors have declared no competing interest.

### Author Declarations

All studies were under the auspices of the Columbia University IRB "Molecular Genetic Analysis of Obesity and Non-Insulin Dependent Diabetes Mellitus" IRB #: AAAA4485 which expires on 5/1/23.

## Bibliography

1. Ward ZJ, Bleich SN, Cradock AL, Barrett JL, Giles CM, Flax C, et al. Projected U.S. State-Level Prevalence of Adult Obesity and Severe Obesity. N Engl J Med. 2019;381(25):2440–50.

2. National Health and Nutrition Examination Survey 2017–March 2020 Prepandemic Data Files Development of Files and Prevalence Estimates for Selected Health Outcomes, (2021).

3. Llewellyn CH, Trzaskowski M, Plomin R, Wardle J. Finding the missing heritability in pediatric obesity: the contribution of genome-wide complex trait analysis. Int J Obesity. 2013;37(11):1506–9.

4. Loos RJF, Janssens Acjw. Predicting Polygenic Obesity Using Genetic Information. Cell Metab. 2017;25(3):535–43.

5. Luke A, Guo X, Adeyemo AA, Wilks R, Forrester T, Lowe W, et al. Heritability of obesity-related traits among Nigerians, Jamaicans and US black people. Int J Obesity. 2001;25(7):1034–41.

6. Maes HHM, Neale MC, Eaves LJ. Genetic and environmental factors in relative body weight and human adiposity. Behav Genet. 1997;27(4):325–51.

7. Albuquerque D, Nobrega C, Manco L, Padez C. The contribution of genetics and environment to obesity. Brit Med Bull. 2017;123(1):159–73.

8. Wang K, Li WD, Zhang CK, Wang Z, Glessner JT, Grant SF, et al. A genome-wide association study on obesity and obesity-related traits. PLoS One. 2011;6(4):e18939.

9. Speakman JR, Loos RJF, O ‘Rahilly S, Hirschhorn JN, Allison DB. GWAS for BMI: a treasure trove of fundamental insights into the genetic basis of obesity. Int J Obes (Lond). 2018;42(8):1524–31.

10. Scuteri A, Sanna S, Chen WM, Uda M, Albai G, Strait J, et al. Genome-wide association scan shows genetic variants in the FTO gene are associated with obesity-related traits. PLoS Genet. 2007;3(7):e115.

11. Khera AV, Chaffin M, Wade KH, Zahid S, Brancale J, Xia R, et al. Polygenic Prediction of Weight and Obesity Trajectories from Birth to Adulthood. Cell. 2019;177(3):587–96 e9.

12. Loos RJF, Yeo GSH. The genetics of obesity: from discovery to biology. Nat Rev Genet. 2022;23(2):120–33.

13. Loos RJ. The genetics of adiposity. Curr Opin Genet Dev. 2018;50:86–95.

14. Marouli E, Graff M, Medina-Gomez C, Lo KS, Wood AR, Kjaer TR, et al. Rare and low-frequency coding variants alter human adult height. Nature. 2017;542(7640):186–90.

15. Lanktree MB, Guo YR, Murtaza M, Glessner JT, Bailey SD, Onland-Moret NC, et al. Meta-analysis of Dense Genecentric Association Studies Reveals Common and Uncommon Variants Associated with Height. Am J Hum Genet. 2011;88(1):6–18.

16. Trynka G, Hunt KA, Bockett NA, Romanos J, Mistry V, Szperl A, et al. Dense genotyping identifies and localizes multiple common and rare variant association signals in celiac disease. Nat Genet. 2011;43(12):1193–U45.

17. Stitziel NO, Peloso GM, Abifadel M, Cefalu AB, Fouchier S, Motazacker MM, et al. Exome Sequencing in Suspected Monogenic Dyslipidemias. Circ-Cardiovasc Gene. 2015;8(2):343-+.

18. Goodrich JK, Singer-Berk M, Son R, Sveden A, Wood J, England E, et al. Determinants of penetrance and variable expressivity in monogenic metabolic conditions across 77,184 exomes. Nat Commun. 2021;12(1).

19. Wilding JPH, Calanna S, Kushner RF. Once-Weekly Semaglutide in Adults with Overweight or Obesity. Reply. N Engl J Med. 2021;385(1):e4.

20. Musunuru K, Pirruccello JP, Do R, Peloso GM, Guiducci C, Sougnez C, et al. Exome sequencing, ANGPTL3 mutations, and familial combined hypolipidemia. N Engl J Med. 2010;363(23):2220–7.

21. Gill R, Cheung YH, Shen YF, Lanzano P, Mirza NM, Ten S, et al. Whole-Exome Sequencing Identifies Novel LEPR Mutations in Individuals with Severe Early Onset Obesity. Obesity. 2014;22(2):576–84.

22. Li P, Tiwari HK, Lin WY, Allison DB, Chung WK, Leibel RL, et al. Genetic association analysis of 30 genes related to obesity in a European American population. Int J Obes (Lond). 2014;38(5):724–9.

23. Feliciano P, Daniels AM, Snyder LG, Beaumont A, Camba A, Esler A, et al. SPARK: A US Cohort of 50,000 Families to Accelerate Autism Research. Neuron. 2018;97(3):488–93.

24. Backman JD, Li AH, Marcketta A, Sun D, Mbatchou J, Kessler MD, et al. Exome sequencing and analysis of 454,787 UK Biobank participants. Nature. 2021;599(7886):628–34.

25. Li H, Ruan J, Durbin R. Mapping short DNA sequencing reads and calling variants using mapping quality scores. Genome Res. 2008;18(11):1851–8.

26. DePristo MA, Banks E, Poplin R, Garimella KV, Maguire JR, Hartl C, et al. A framework for variation discovery and genotyping using next-generation DNA sequencing data. Nat Genet. 2011;43(5):491–8.

27. Pedersen BS, Quinlan AR. Who ‘s Who? Detecting and Resolving Sample Anomalies in Human DNA Sequencing Studies with Peddy. Am J Hum Genet. 2017;100(3):406–13.

28. Manichaikul A, Mychaleckyj JC, Rich SS, Daly K, Sale M, Chen WM. Robust relationship inference in genome-wide association studies. Bioinformatics. 2010;26(22):2867–73.

29. McLaren W, Gil L, Hunt SE, Riat HS, Ritchie GRS, Thormann A, et al. The Ensembl Variant Effect Predictor. Genome Biol. 2016;17.

30. Wang K, Li MY, Hakonarson H. ANNOVAR: functional annotation of genetic variants from high-throughput sequencing data. Nucleic Acids Res. 2010;38(16).

31. Karczewski KJ, Francioli LC, Tiao G, Cummings BB, Alfoldi J, Wang Q, et al. The mutational constraint spectrum quantified from variation in 141,456 humans. Nature. 2020;581(7809):434–43.

32. Ioannidis NM, Rothstein JH, Pejaver V, Middha S, McDonnell SK, Baheti S, et al. REVEL: An Ensemble Method for Predicting the Pathogenicity of Rare Missense Variants. Am J Hum Genet. 2016;99(4):877–85.

33. Poplin R, Chang PC, Alexander D, Schwartz S, Colthurst T, Ku A, et al. A universal SNP and small-indel variant caller using deep neural networks. Nat Biotechnol. 2018;36(10):983-+.

34. Yun T, Li H, Chang PC, Lin MF, Carroll A, McLean CY. tAccurate, scalable cohort variant calls using DeepVariant and GLnexus. Bioinformatics. 2020;36(24):5582–9.

35. Bycroft C, Freeman C, Petkova D, Band G, Elliott LT, Sharp K, et al. The UK Biobank resource with deep phenotyping and genomic data. Nature. 2018;562(7726):203-+.

36. Mbatchou J, Barnard L, Backman J, Marcketta A, Kosmicki JA, Ziyatdinov A, et al. Computationally efficient whole-genome regression for quantitative and binary traits. Nat Genet. 2021;53(7):1097-+.

37. De Rosa MC, Glover HJ, Stratigopoulos G, LeDuc CA, Su Q, Shen YF, et al. Gene expression atlas of energy balance brain regions. Jci Insight. 2021;6(16).

38. Zhai R, Olias G, Chung WJ, Lester RAJ, Dieck ST, Langnaese K, et al. Temporal appearance of the presynaptic cytomatrix protein bassoon during synaptogenesis. Mol Cell Neurosci. 2000;15(5):417–28.

39. Schoch S, Gundelfinger ED. Molecular organization of the presynaptic active zone. Cell Tissue Res. 2006;326(2):379–91.

40. Altrock WD, Dieck ST, Sokolov M, Meyer AC, Sigler A, Brakebusch C, et al. Functional inactivation of a fraction of excitatory synapses in mice deficient for the active zone protein bassoon. Neuron. 2003;37(5):787–800.

41. Angenstein F, Niessen HG, Goldschmidt J, Lison H, Altrock WD, Gundelfinger ED, et al. Manganese-enhanced MRI reveals structural and functional changes in the cortex of bassoon mutant mice. Cereb Cortex. 2007;17(1):28–36.

42. Gundelfinger ED, Reissner C, Garner CC. Role of Bassoon and Piccolo in Assembly and Molecular Organization of the Active Zone. Front Synaptic Neurosci. 2015;7:19.

43. Hallermann S, Fejtova A, Schmidt H, Weyhersmuller A, Silver RA, Gundelfinger ED, et al. Bassoon Speeds Vesicle Reloading at a Central Excitatory Synapse. Neuron. 2010;68(4):710–23.

44. Waites CL, Leal-Ortiz SA, Okerlund N, Dalke H, Fejtova A, Altrock WD, et al. Bassoon and Piccolo maintain synapse integrity by regulating protein ubiquitination and degradation. Embo J. 2013;32(7):954–69.

45. Hashida H, Goto J, Zhao ND, Takahashi N, Hirai M, Kanazawa I, et al. Cloning and mapping of ZNF231, a novel brain-specific gene encoding neuronal double zinc finger protein whose expression is enhanced in a neurodegenerative disorder, multiple system atrophy (MSA). Genomics. 1998;54(1):50–8.

46. Montenegro-Venegas C, Annamneedi A, Hoffmann-Conaway S, Gundelfinger ED, Garner CC. BSN (bassoon) and PRKN/parkin in concert control presynaptic vesicle autophagy. Autophagy. 2020;16(9):1732–3.

47. Yabe I, Yaguchi H, Kato Y, Miki Y, Takahashi H, Tanikawa S, et al. Mutations in bassoon in individuals with familial and sporadic progressive supranuclear palsy-like syndrome. Sci Rep-Uk. 2018;8.

48. Zheng H, Berthoud HR. Neural systems controlling the drive to eat: mind versus metabolism. Physiology (Bethesda). 2008;23:75–83.

49. Shah BP, Vong L, Olson DP, Koda S, Krashes MJ, Ye CP, et al. MC4R-expressing glutamatergic neurons in the paraventricular hypothalamus regulate feeding and are synaptically connected to the parabrachial nucleus. P Natl Acad Sci USA. 2014;111(36):13193–8.

50. Fenselau H, Campbell JN, Verstegen AMJ, Madara JC, Xu J, Shah BP, et al. A rapidly acting glutamatergic ARC -> PVH satiety circuit postsynaptically regulated by alpha-MSH. Nat Neurosci. 2017;20(1):42–51.

51. Claflin KE, Sullivan AI, Naber MC, Flippo KH, Morgan DA, Neff TJ, et al. Pharmacological FGF21 signals to glutamatergic neurons to enhance leptin action and lower body weight during obesity. Mol Metab. 2022;64:101564.

