## Supplemental figures for "Predicted loss of function alleles in Bassoon (BSN) are associated with obesity"

Supplementary Figure 1. QQ-plots of Columbia data vs SPARK cohort

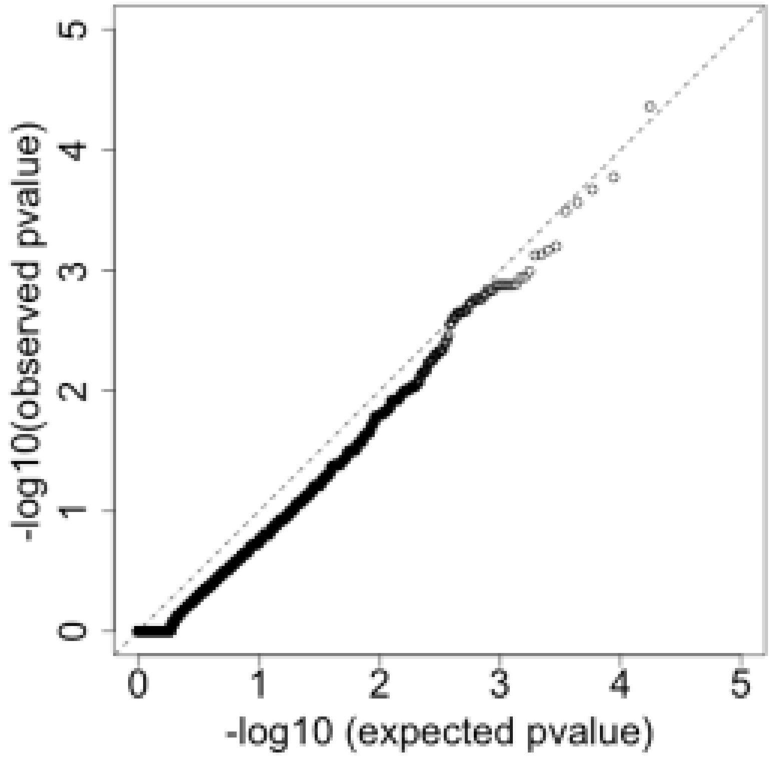


Syn, lambda=0.75

Supplementary Figure 1.

QQ plots comparing observed pvalue to expected pvalue for the UKBB data. A.) QQ-plot for synonymous variants demonstrating no inflation since lambda = 1. B.) QQ-plot for the gene-based association test for likely gene damaging and deleterious variants (using revel >=0.55 as threshold), indicating some deflation (lambda=0.93).

Supplementary Figure 2. QQ-plot of UKBB data with synonymous variants, and LGD+REVEL0.55

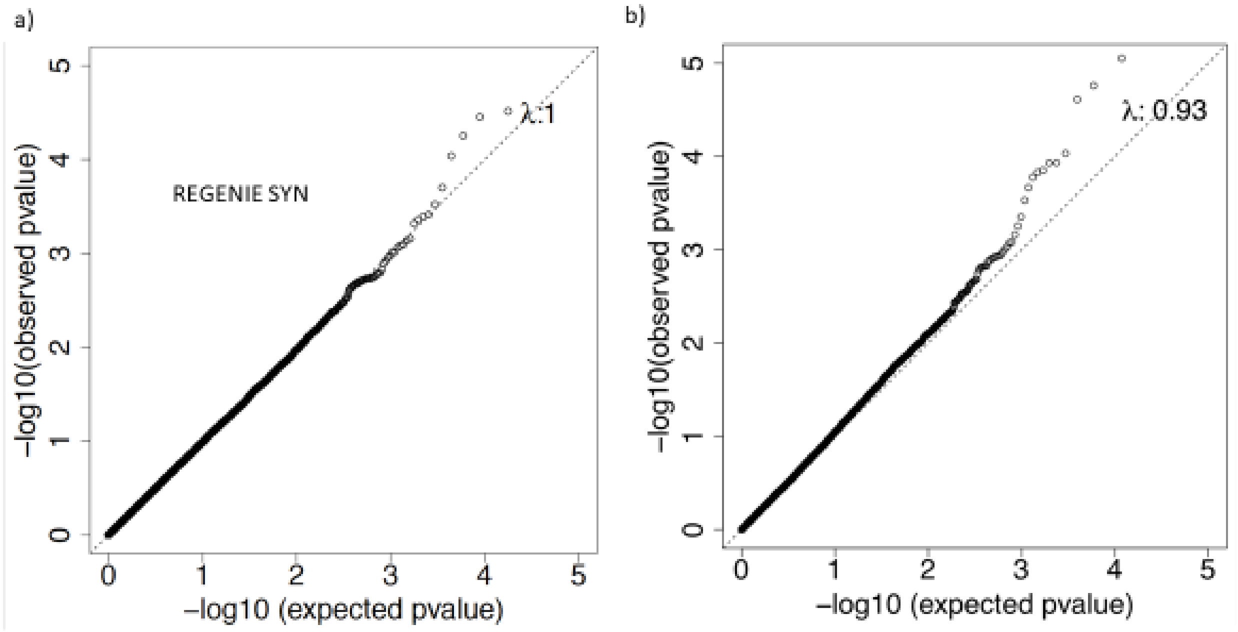


Supplementary Figure 2.

QQ plots comparing observed pvalue to expected pvalue for the Columbia data (using the SPARK data as controls). A. QQ-plot for synonymous variants, lambda is .75, attributable to the small sample size causing a step to the right from missing genes with synonymous variants. B.) QQ-plot for the gene-based association test for pLOF and Dmis (using revel >=0.55 as threshold), indicating some deflation (lambda=0.66). Once again, the curve is stepped to the right caused by the small sample size in the cohort.

Supplemental Figure 3.

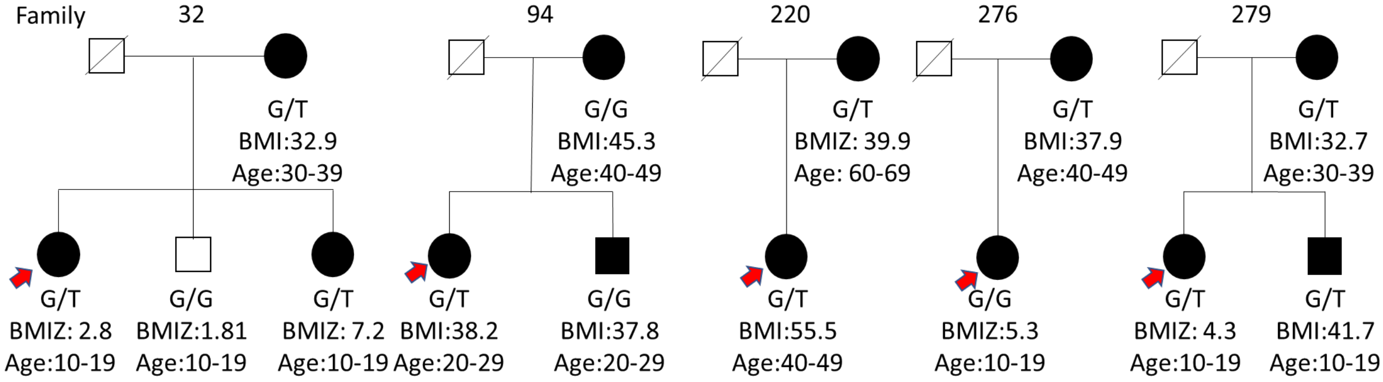


6_10671528_10671528_G_T (rs887287256)

C6orf52 Segregation

All carriers from the same batch, VCR;

Supplementary Figure 3.

Segregation analysis in the Columbia cohort for rs88728256 which reached genome-wide significance. Filled in symbols indicates obese individuals, red arrow indicates proband. All heterozygotes are affected, but not affected individuals are heterozygous for this variants.

**Supplemental Figure 4**

**CTRC 1_15445606_15445606_G_A**

Rs202058123

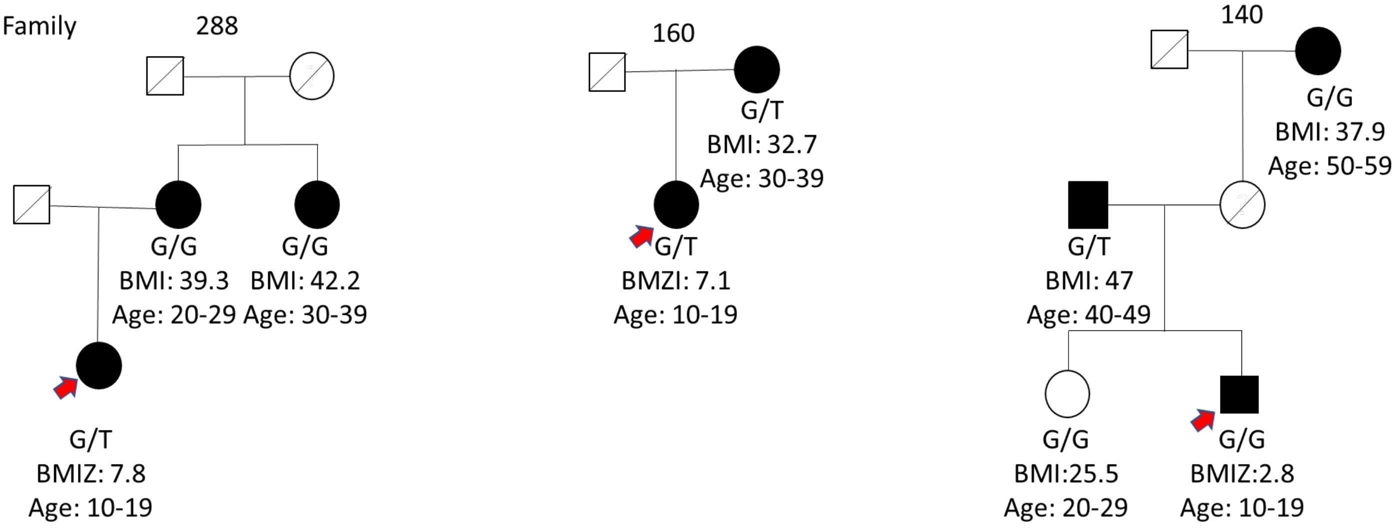


Supplementary Figure 4.

Pedigree for variant rs202058123 (*CTRC* p.G217S) from the Columbia cohort. Filled in symbols indicate obese, red arrows indicate proband. All heterozygotes are affected, but not affected are heterozygous.

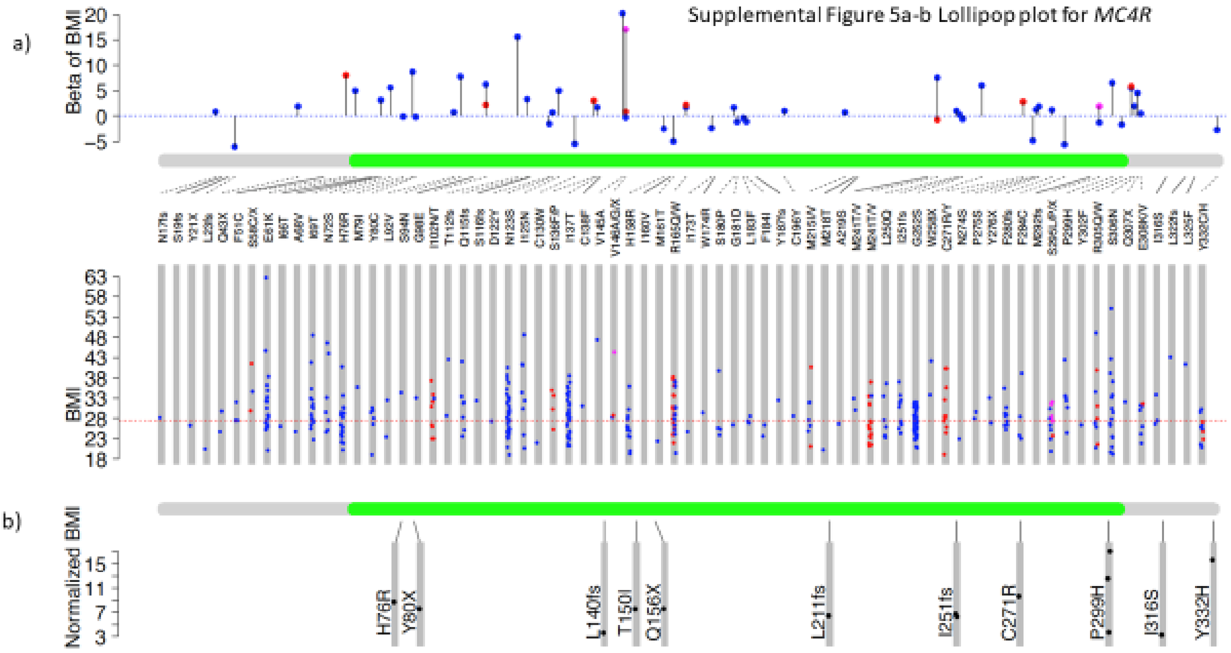


Supplementary Figure 5.

Lollipop plot of BMI for *MC4R* variants in UKBB cohort. Amino acid variants at the same position were sorted alphabetically, corresponding to the color order: blue, green, magenta, orange. Colors are matched in the upper and lower panels. The dashed line indicated if the variant is included in the REGENIE single variant result.

The full list of variants in UKBB and Columbia cohort are listed in the table file.

Supplemental Figure 6 BMI in *MC4R* and *MEOX1* heterozygotes.

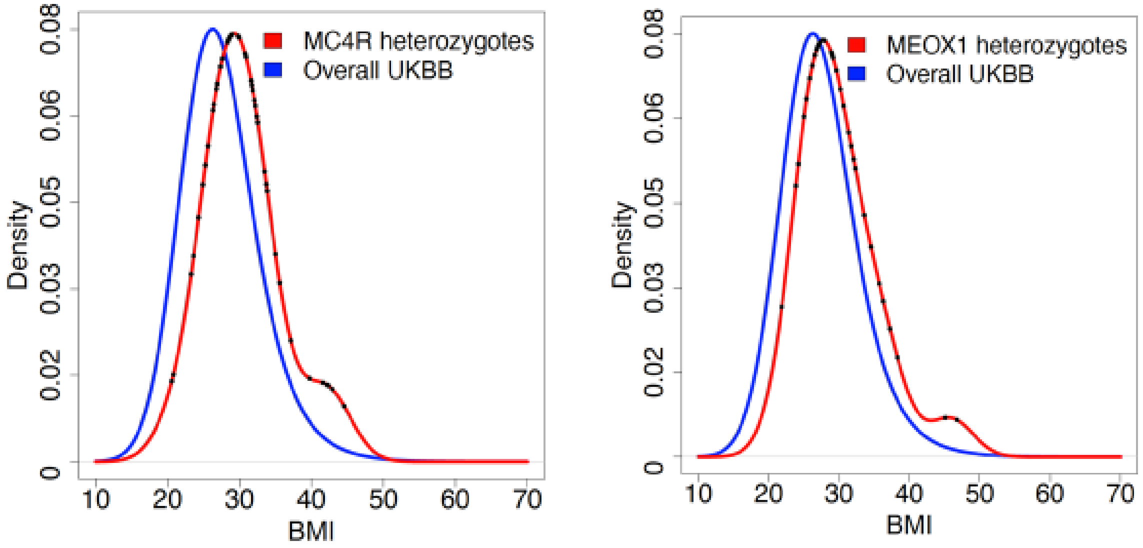


Supplementary Figure 6.

BMI density distribution for the heterozygous *MC4R* pLoF variants and heterozygous *MEOX1* Dmis and pLoF individuals. Both BMI distributions are shifted to the right in the heterozygotes.
